# Comparison of the Vaginal and Labial Dimensions of Nullipara Ethnic Chinese and Western Women

**DOI:** 10.1101/2024.05.18.24307569

**Authors:** Lisa Stevens, Mariana Masteling, Kruthi Srinivasa Raju, Sara Mastrovito, James A. Ashton-Miller, John O. L. DeLancey

## Abstract

**Introduction and Hypothesis:** Vaginal dimensions have clinical and surgical implications. We sought to quantify the differences between vaginal and labial dimensions in healthy ethnic Chinese and Western women with normal pelvic organ support.

**Methods:** This is a cross-sectional study of a convenience sample of ethnic Chinese nullipara (n = 33) and Western nullipara (n= 33) women recruited for research purposes. For each subject, magnetic resonance imaging was used to quantify the vaginal and labial dimensions. Specifically, we identified the anterior and posterior vaginal wall, the outline of the cervix in the mid-sagittal and coronal planes, and the distance from the labia majora to the hymenal ring at the urethral meatus.

**Results:** The vaginal and labial dimensions of ethnic Chinese nullipara were 9-21 % smaller than those of Western nullipara; however, the relative orientation of the cervix to the vaginal opening was not different.

**Conclusions:** Significant group differences in vaginal and labial dimensions were found, with Chinese nullipara dimensions being up to 21% smaller than those of Western nullipara.

**Brief Summary:** Ethnic Chinese women have smaller vaginal and labial dimensions than Western women. These differences have clinical and surgical implications.

## Introduction

Studies of female pelvic floor anatomy and disorders often have mixed ethnic group representation, rather than data from a single group. Given the increasing globalization of treatment recommendations, consensus statements, and medical products, considering how members of various ethnic groups might differ from each other can affect the optimal care for each group. Prior studies of ethnic group differences have demonstrated differences in cervical length [1], pelvic bony anatomy [2,3], the risk for perineal tear during birth [4], and prevalence of pelvic organ prolapse [5]. However, to our knowledge, quantitative ethnic or racial differences in vaginal and labial dimensions between Western and ethnic Chinese women have not been measured.

There is a general lack of data on the variation in vaginal and labial dimensions of healthy women. For example, while studies have used endovaginal casts to observe the range of vaginal shapes, this method is not easily scalable and replicated [6]. A 2006 magnetic resonance imaging (MRI) study of vaginal dimensions of 28 Western women provided baseline measurements [7], and a 2016 study employed quantitative MRI analysis methods to measure selected vaginal dimensions in 80 Western women [8].

Ethnic and racial differences in vaginal and labial dimensions have implications for clinical care, medication dosing, fit of intravaginal devices such as pessaries and contraceptive devices, and design of surgical products. These dimensions can also be affected by parity. Ethnic Chinese women are a large and historically understudied population regarding vaginal dimensions. We therefore conducted a cross-sectional study to test the hypothesis that there are no differences in vaginal and labial dimensions between ethnic Chinese and Western nullipara as measured on MRI.

## Materials and Methods

### Datasets

#### Ethnic Chinese Group

We defined ethnic Chinese women as having both parents from either China or Taiwan, and both maternal and paternal grandparents being from mainland China. Thirty-three healthy premenopausal nulliparous adult ethnic Chinese women were recruited from southeast Michigan, United States. Each individual underwent pelvic MRI scans for research purposes between March and June 2021. Exclusion criteria included current pregnancy, previous hysterectomy or pelvic floor surgery, radiation to the pelvis, intravaginal scarring, active vaginal infection, vaginismus, or fistulas. This study received approval from our Institutional Review Board and all subjects gave written consent to participate.

Each subject underwent MRI while resting in the supine position. Ultrasound gel was placed into the vagina to improve visualization if the subject consented. Proton density-weighted fast spin-echo imaging was performed in the axial, sagittal, and coronal planes using a 3-Tesla (3-T) Ingenia MRI scanner (Philips Medical Systems, Best, The Netherlands) with 2 mm slice thickness and a 1 mm gap between slices.

#### Western Nulliparous Group

We selected a convenience sample of 33 MRI scans of Western nullipara from our institution’s pelvic floor MRI repository that were age-matched to the Chinese sample [9–11]. Women were considered Western if they did not identify as Asian.

### Vaginal and labial dimensions quantification

MR image analysis was performed in 3D Slicer [12] using a technique consistent with a prior study [8]. In the sagittal plane, fiducials (points) were placed to identify the anterior and posterior vaginal wall and cervix (Figure 1a). When gel was present in the vagina, the mid-vaginal line was also identified. Using these markers, the distances from the vaginal opening to the anterior fornix, the vaginal opening to the cervix, and the vaginal opening to the posterior fornix were measured. The level of the vaginal opening (introitus) was established at the distal end of the urethra, which lies at the level of the hymenal ring in the coronal plane. Using the anterior wall fiducial list, the vaginal wall was divided into two segments in order to obtain the vaginal axis, with the mid-vaginal point defined as the point closest to the half-vertical distance between the vaginal opening and the anterior fornix. The cervical axis was defined by the anterior and posterior vaginal fornix and represented a third axis.

**Figure 1.**
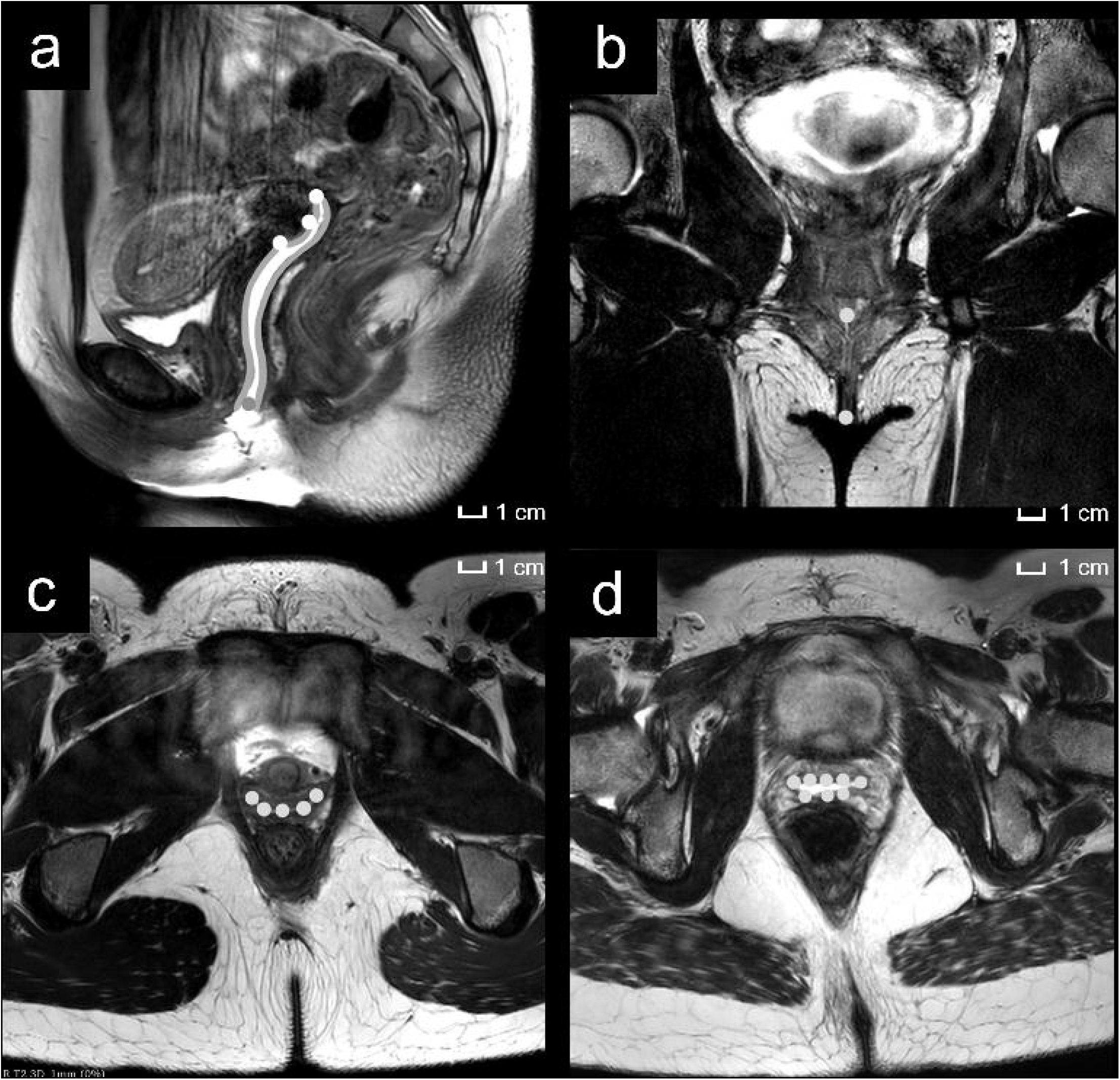
MRI Measurements. a: Examples of fiducials placed in the sagittal plane outlining the anterior and posterior vaginal walls and mid-vaginal line in a Chinese nullipara subject with gel. b: Example of the labial distance fiducials placed in the coronal plane in an ethnic Chinese woman. c, d: Five or eight fiducials, respectively, placed axially in the vagina in order to trace the perimeter of the vagina (Chinese nullipara).

In the axial plane, in slices separated by 5 mm, fiducials were placed on the vaginal wall between the introitus and the anterior fornix in order to identify the anterior and posterior vaginal walls (Figure 1b, 1c). The vaginal width was measured as the distance between the two most lateral points. To quantify vaginal width differences, the length of the anterior vaginal wall was divided into four segments and the mean ± standard deviation (SD) was calculated for each group.

The distance from the labia majora to the vaginal opening was measured in the coronal plane in the slice where the introitus was visible, with a fiducial placed at the point where the labia majora meet. The distance from that point to the level of the hymen as the external urethral meatus was measured as the ‘labial distance’ (Figure 1d). All points were normalized to the Pelvic Inclination Correction system (PICS) [13].

### Data and statistical analysis

Data management and statistical analysis were performed in Python (Python Software Foundation, Wilmington, DE USA). Demographics and descriptive statistics (mean ± SD) of vaginal and labial dimensions were calculated for both groups. If dimensions were statistically different, the percentage difference was also calculated as the ratio of the difference of group means divided by the mean of the Western nullipara. The angle and distance between the vaginal opening and mid-vaginal point, between the mid-vaginal point and the anterior fornix, and between the anterior fornix and the posterior fornix were calculated for each subject. Population percentiles were also calculated for each group.

Tests for normality were conducted using the Python SciPy library based on D’Agostino and Pearson’s methodology [14]. Two-sided Independent Student’s t-tests were performed using a significance level of 0.05. Spearman and Pearson’s correlations were used to investigate linear and monotonic relationships between vaginal and labial dimensions and demographics; only correlations with p<0.05 and correlation coefficient r≥0.5 are reported.

## Results

Thirty-three ethnic Chinese nulliparous and 33 Western nulliparous were included for analysis (Table 1). Age and weight were significantly different between nulliparous groups (p<0.001, p<0.05). Substantial variation in vaginal and labial dimensions was found within each group. The vaginal and labial dimensions of ethnic Chinese nullipara ranged from 9-21 % smaller than those of Western nullipara (Table 1); the angle between the cervix and the PICS line was similar between groups (Figure 2, Table 1).

**Table 1.**
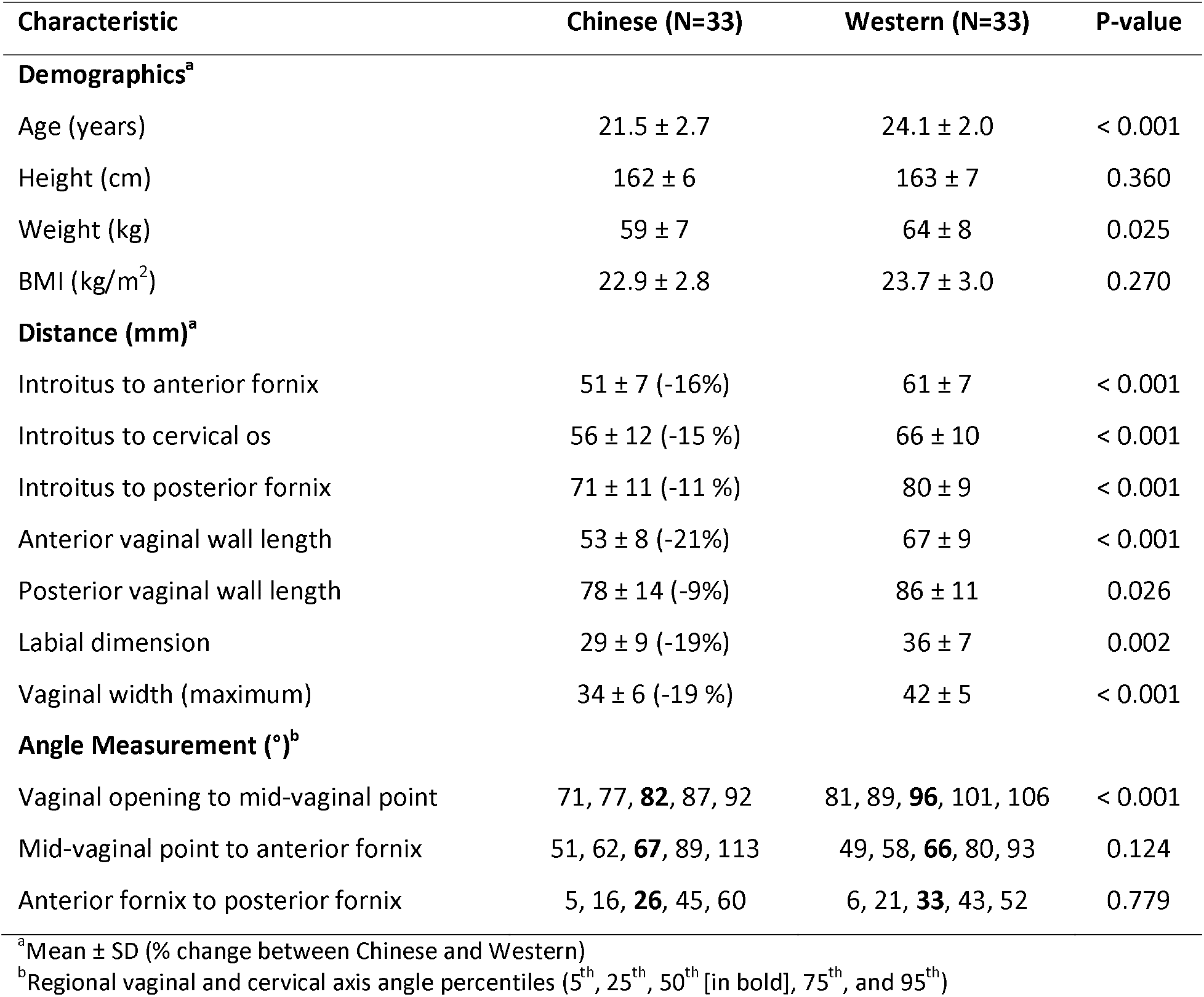
Comparison of demographic data and vaginal and labial dimensions between ethnic Chinese and Western nullipara

**Figure 2.**
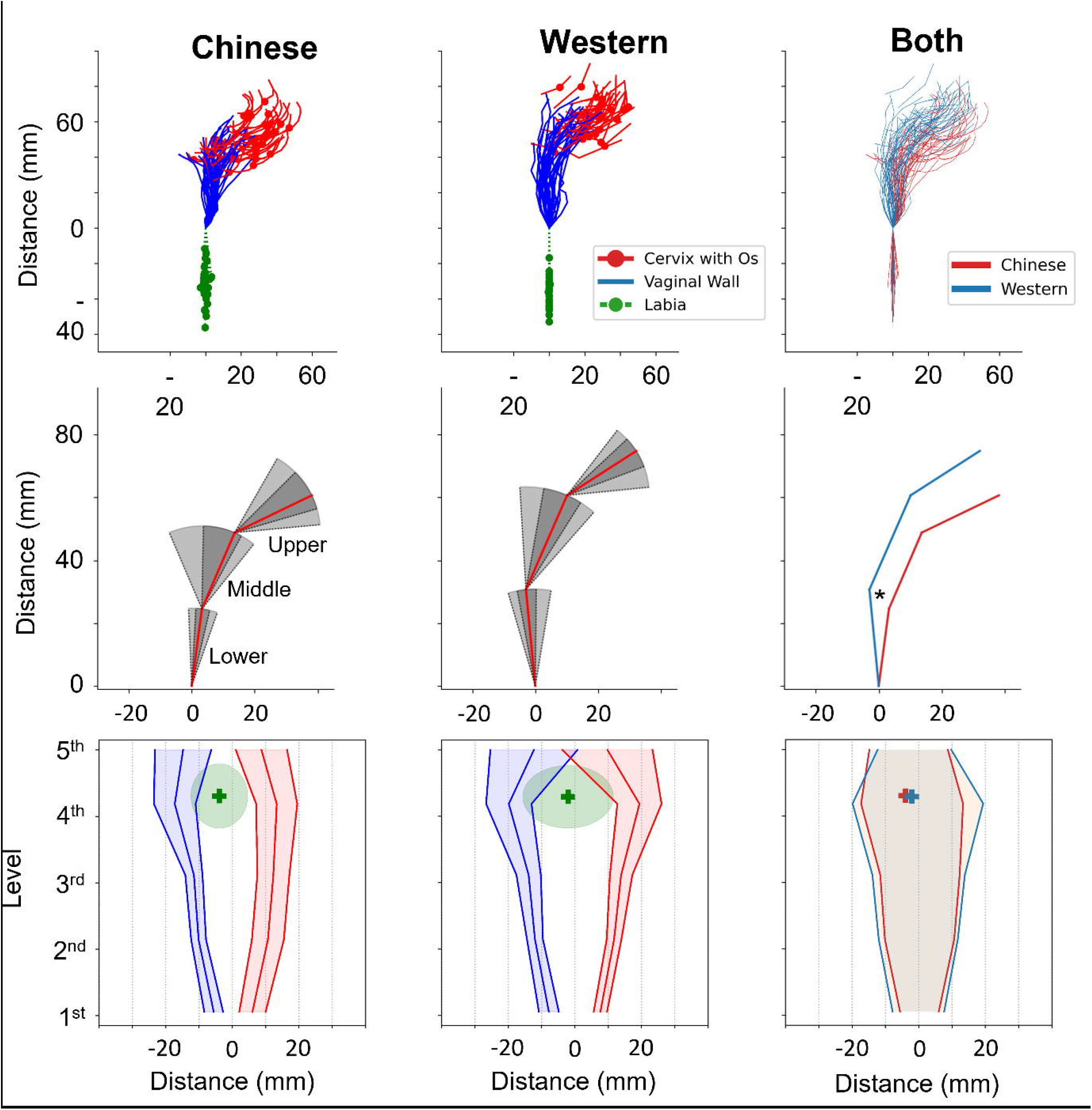
Ethnic Chinese versus Western nullipara vaginal and labial dimensions. Top row: Graphic display of the distribution of the location and orientation of the cervix (red), cervical os (red circle), vaginal wall (blue), and labial distance (green) for both groups. The graph at the far right represents the mean of all subjects superimposed, with Chinese nullipara in red and Western nullipara in blue. Middle row: 5^th^, 25^th^,50^th^, 75^th^ and 95^th^ percentile sagittal outlines of the anterior vaginal wall divided in two segments and the cervix defined by the anterior and posterior fornix points. Percentile values are reported in Table 1. The graph on the far right shows the 50^th^ percentile for both groups, with the * indicating the only statistically significant portion (angle of segment between vaginal opening to mid-vaginal point). Bottom row: Coronal view of the mean ± SD of the location of the left (blue) and right (red) vaginal wall widths, along with the cervix location (green). Vaginal widths for each level are reported in Table 1.

In the ethnic Chinese group, increasing weight and BMI correlated with greater labial distance (r= 0.66 and r = 0.63, respectively), as did the distance from the vaginal opening to the cervical os (r=0.5). In the Western group, only the weight correlated with the labial distance (r = 0.51).

## Discussion

Ethnic Chinese vaginal and labial dimensions were systematically smaller than those of Western nulliparous women. The effect of age and parity on ethnic Chinese vaginal and labial dimensions is unknown.

There are several strengths to this work. It compares the vaginal dimensions of two ethnic groups acquired specifically for research purposes, rather than using scans of individuals presenting to the clinic with clinical problems that might have biased the results. It employed high-resolution MRI specifically designed for showing the soft tissues of the pelvis and involved minimal distortion of the vagina compared to techniques involving a molding compound [6] or the presence or pressure of an ultrasound probe. This work also expands the findings of Luo et al [8] by adding labial distances. By quantifying the differences between ethnic Chinese and Western nullipara, we can begin to address this knowledge gap and thereby ensure that women can receive care designed specifically for them, rather than universal care based on the Western population.

Limitations should be kept in mind while interpreting our results. The ethnic Chinese sample size was modest and the use of convenience sampling for the Western group might have included a selection bias toward clear MR images. While the fact that a limited amount of ultrasound gel was inserted into the vagina of some subjects in each group could have affected the measurements slightly, it improved the definition of the vaginal margins. There were small but statistically significant differences in the ages of the ethnic Chinese and Western nulliparous groups (24.1 vs. 21.5 years, p<0.001) and in their weights (59 ± 7 vs. 64 ± 8 kg, p<0.05), but without differences in BMI, this probably had little practical impact, as both groups were of similar age and body constitution. Given the many ethnic groups and large population in China, the present sample is unlikely to be representative of the Chinese population. Next steps could include taking measurements of women from the various Chinese ethnic groups, as well as of women of other races and ethnicities. This would provide quantification and a more well-rounded view of the variations in the vaginal and labial dimensions across ethnic and racial groups.

This study provides new information on differences in vaginal anatomy between two different ethnic groups. It extends existing studies that report racial differences in internal anatomy such as levator hiatal area and pubovisceral muscle thickness [15,16], cervical length [1], pelvic organ prolapse [5,17], pelvic anatomy [2,3], and risk for perineal laceration during birth [4,18].

The smaller vaginal and labial dimensions found in Chinese nullipara compared to Western nullipara are important for clinical and surgical planning. With the globalization of medical literature, products, and standardized staging systems and recommendations, recognizing differences in anatomy among different ethnic groups can help ensure each group receives treatment and recommendations best suited to them. Further, with obesity increasing rapidly across the world, a better understanding of its effects on external female anatomy is necessary, despite its lack of effect on internal vaginal dimensions.

In conclusion, the vaginal and labial dimensions of Chinese nullipara were up to 21% smaller than those of Western nullipara. This finding is important for clinical and surgical planning, as well as for device design.

## Data Availability

All data produced in the present study are available upon reasonable request to the authors

## Notes

**Financial disclaimer/Conflict of interest:** None

### Competing Interest Statement

The authors have declared no competing interest.

### Funding Statement

This study was funded by a research grant from Procter & Gamble and the National Institutes of Health under grants R01 HD038665, P50 HD044406, and RC2 DK122379 grants.

### Author Declarations

IRB of University of Michigan have ethical approval for this work

